# Measuring the Impact of Global R&D Investment in Product Development Partnerships (PDPs): A Case Study on Return on Investment in Antimalarial Drug Development

**DOI:** 10.1101/2025.06.17.25329737

**Authors:** Paula Christen, Céline Audibert, Jo-Ann Mulligan, Oliver Bubb-Humfryes, Christian von Drehle, Lesong Conteh

## Abstract

**Background:** Product Development Partnerships (PDPs) are non-profit organizations that bridge the gap between the need for new treatments for poverty-related diseases and the resources available to develop them, leveraging a mix of public, philanthropic, multilateral, and private sector funding. This paper describes how two financial metrics were used to estimate the return on investments of drugs developed by the PDP Medicines for Malaria Venture (MMV) as a case study.

**Methods:** The internal rate of return (IRR) and benefit-cost ratio (BCR) were used to estimate the economic return on investment for the PDP. IRR was based on total investments from 2000 to 2023 and health gains derived from PDP-supported drugs, measured as monetized disability-adjusted life years (DALYs) minus the delivery cost of the products. BCR was calculated by dividing the present value of monetized DALYs by the present value of cost, indicating the overall efficiency and impact of the investments received by MMV.

**Findings:** Total investment received was $2·3 billion over the study period, and the antimalarial drugs developed and launched with the support of MMV averted an estimated 1·6 million deaths and 87 million DALYs for a cost of delivery estimated at $785 million. The IRR for the base scenario was 52·13% (CI: 52·11% - 52·16%) and the BCR 12·99 (CI: 12·92 - 13·06).

**Interpretation:** The substantial IRR and BCR generated by investment in antimalarial drug development suggest that the PDP model has a potentially pivotal role to play in global health.

**Funding:** No funding to declare.

## Research in context

### Evidence before this study

We searched PubMed and manually inspected the results using “internal rate of return”, “IRR”, “benefit-cost ratio*”, “BCR”, “return on investment”, “ROI”, “product development partnerships”, “PDP” as keywords, in English, and up until February 2025. We also performed a search in the grey literature and included in our review reports and articles published by consulting organizations, funders, government organisations, and development banks.

Both internal rate of return (IRR) and benefit-cost ratio (BCR) are widely used to inform investment decisions in the public and private sector. The need for better data to inform global health R&D investments is long recognised and our searches indicate a continued lack of consensus on the preferred metrics and methods.^1^ A recent scoping review showed that there were inconsistencies in the calculation and interpretation of ROI associated with public health interventions.^1^ The authors called for more transparency when reporting underlying assumptions.

Most economic evaluations focused on low- and middle-income countries’ estimate the cost-effectiveness of specific interventions or products.^2^ Literature on the benefit cost ratios and estimates on the value of global health investments in monetary terms are uncommon.^3,4^ Moreover, understanding the return on investing in global R&D is rarer still.^5^ In the UK, a series of influential analyses using the Internal Rate of Return (IRR) demonstrated the economic returns of medical research funding for four diseases (cancer, mental health, cardiovascular diseases, and musculoskeletal diseases).^6^ To date, no peer-reviewed publications have evaluated the return on investment in the R&D generated by product-development partnerships (PDPs).

### Added value of this study

This paper contributed to knowledge in several ways. Firstly, from an empirical perspective we learn that investing in malaria research and development through the PDP model has resulted in a positive return on investment, driven by providing access to effective antimalarial treatments, averting 1·6 million deaths. Secondly, we demonstrate that a rigorous analysis of PDP returns on investment can be conducted using routine data and standardised methods to inform both productive and allocative efficiency decisions. Finally, from a policy perspective, this paper is able to show that investment in R&D, and PDPs specifically, are a competitive investment when compared to other health and non-health investments.

### Implications of all the available evidence

In a funding environment where large-scale investments are increasingly coming under scrutiny, this work demonstrates that it is possible to calculate the return on investment from PDPs using routine data and standardised methods. This paper offers a retrospective and conservative view of what has been achieved. Even with this more conservative methodology, it is clear that the investment received by MMV allowed it to respond to existing market failures and develop a portfolio of drugs that have positive health and economic returns. Our paper is an example of how global health investments can assess their returns on research investment in a transparent and robust way to inform policy making and funding allocation.

## INTRODUCTION

Product Development Partnership (PDPs) are non-profit organizations that bridge the gap between public health needs and market-driven incentives by coordinating and financing the research, development, and delivery of new health products for diseases of poverty.^7–9^ They were developed to address market failures, specifically, the gap between the unmet need for new treatments and interventions for poverty-related and neglected diseases in low- and middle-income countries, and the resources available to develop them.^10^ PDPs are non-profit organisations that derisk the product development process by bringing together partners including pharmaceutical companies, research organisations, academic institutions, multilateral agencies, and other stakeholders such as user and advocacy groups.^7,8^ The funding of PDPs is based on a mix of resources combining public sector funding from governments, philanthropic foundations such as the Gates Foundation, multilateral organizations, and the private sector with pharmaceutical and biotechnology companies providing various kinds of contributions.^11^

Since 2010, a coalition of twelve leading PDPs have delivered nearly 80 new health technologies reaching over 2·4 billion people.^10^ Quantifying the impact of funding PDPs in the context of new health technology development and increased access is essential. Additionally, understanding the return on investing in PDPs in monetary terms enables comparisons with other potential uses of funding for global health and development more broadly.^1^ Direct commercial returns or profit are not a marker of success for PDPs; returns on investment must therefore be demonstrated using approaches that extend beyond cash flows. These commonly include health metrics such as cases averted, lives saved, disability adjusted life years (DALYs) averted, quality adjusted life years (QALYs) gained, or cost-effectiveness estimates.^10–12^

Return on investment estimates often forecast bold and convincing cases for funding global health interventions.^3,4^ The importance of investing in research and development (R&D) has been proven across a range of diseases. A recent high profile modelling analysis forecasting the health economic and social impact of 20 years of investment in global health R&D across multiple diseases showed impressive returns on investments.^5^ This analysis estimated that every US$1 invested in neglected disease R&D generates a return of US$405 by 2040, and that at least 40·7 million lives are to be saved for the period 2000 to 2040, averting 2·83 billion DALYs. The analysis provided strong justification for continued investment across a range of diagnostics and treatments for multiple neglected diseases in global health R&D, it did not however, investigate contributions at the individual PDP level.

In this paper, we estimate the returns from investing in a PDP using two widely accepted financial metrics: the Internal Rate of Return (IRR) and the Benefit-Cost Ratio (BCR).^6,12–14^ Using the PDP Medicines for Malaria Venture (MMV), a not-for-profit organization whose mission is to deliver a portfolio of accessible medicine with the power to treat, prevent, and eliminate malaria, as a case study, we assess the benefits of PDP investments in terms of health and economic gains achieved per dollar invested, while acknowledging potential risks such as inefficient resource use or delayed access to innovations.

## METHODS

### A. Overall approach

The IRR and BCR were used to estimate the efficiency of funding received by MMV over time. Most commonly used in the private sector, IRR values have also informed policy decisions in the public sector. For example, the UK government determined the IRR achieved by medical research in four disease areas to estimate the economic returns of public investment.^6,12^ The BCR is a metric frequently used to estimate the ROI of public health interventions (Table 1).^13,14^

**Table 1:** IRR and BCR characteristics.

| Characteristics | Internal Rate of Return (IRR) | Benefit-Cost Ratio (BCR) |
| --- | --- | --- |
| **Output format** | Percentage (%) | Ratio<br>Example: a BCR of 1.5 means \$1.50 of benefit is generated from every \$1 spent |
| **Focus** | Annualised rate of return over time | Value gained per dollar (unit of currency) |
| **Time consideration** | Considers timing of cash flows | Does not factor in time, focus is total values |
| **Decision criteria** | Compares IRR to a target rate | BCR > 1 indicates a good project |
| **Context most often used** | - Business and Private Sector Projects where the timing of cash flows is an important consideration.<br>- Comparison of projects, often at similar scales. | - Public Sector/Social Projects where benefits are non-monetary and impact is not purely financial.<br>- Accountably to taxpayers and wider stakeholders a driving factor.<br>- Assessment of whether large capital investments are worth undertaking |

Returns from the PDP’s product portfolio were estimated as the monetized value of DALYs averted minus the costs of product delivery in malaria-endemic countries. The steps to estimate the IRR and BCR are described below (Box 1).

#### Box 1: Formula

**IRR calculation**

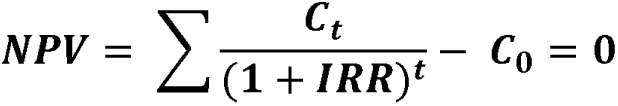

**Where:**

NPV = net present value

C_t_ = net cash inflows during the period t

C_0_ = total initial investment costs

IRR = the internal rate of return

t = the number of periods

The IRR is the discount rate at which the NPV of all cash flows equals zero, that is, the rate that equates the present value of benefits and costs over time.

**BCR calculation**

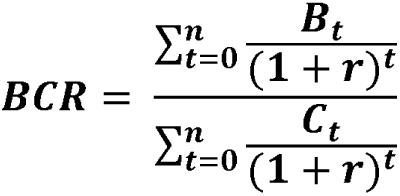

Where:

B = benefits, in our case the monetized DALYs

C = cost of delivery plus cost of investment received by MMV

r = discount rate

*t = year*

*n = analytical horizon (in years)*

### Step 1: Calculating total investment costs

Total investment costs (C_0_) consisted of the funding received by MMV to develop antimalarial drugs for the period 2000 to 2023 (as provided by MMV) and expressed in 2023 US$ prices. Total investment costs included investment for products that did not make it to market, as well as those that were still in development at the time of the analysis and those already launched. Comparing the total funding received by the PDP with the benefits generated by products that successfully reached the market ensures that the analysis accounts for the full cost of the R&D process, including both successful and unsuccessful development efforts. The time periods spanned from the creation of the PDP in 2000 to 2023, the most recent year for which cost of delivering products were available at the time of writing.

### Step 2: Estimating the number of deaths averted

The first step in the calculation of the health gain (the C_t_ component of the equation) is to estimate the number of deaths averted. To achieve this the volumes of the PDP-supported products procured and sold cumulatively since date of market entry or WHO-prequalification were obtained from manufacturers based on routine data collection from MMV. A timeline showing the year of entry of each product is provided in the supplementary material A. This approach accounts for product use in both public and private settings. Volumes are converted into number of treatments provided based on the estimated number of doses required per treatment and for each molecule. This generates a theoretical number of people treated or protected which is further adjusted for wastage of volume sales and appropriate use to obtain the estimated number of people effectively treated or protected.^15^ To estimate the number of deaths averted, the protective effect of each product is compared with the most likely counterfactual. The efficacy parameters used in the model were derived from peer-reviewed literature and systematic reviews used by WHO to update their malaria guidelines as available at the time of the study. Table 2 presents the assumptions used in the model to convert the volumes of each product into the number of cases and deaths averted. The model followed a conservative approach by only estimating the number of deaths averted up until 2023, not taking into account any future deaths averted from PDP supported products.

**Table 2.**
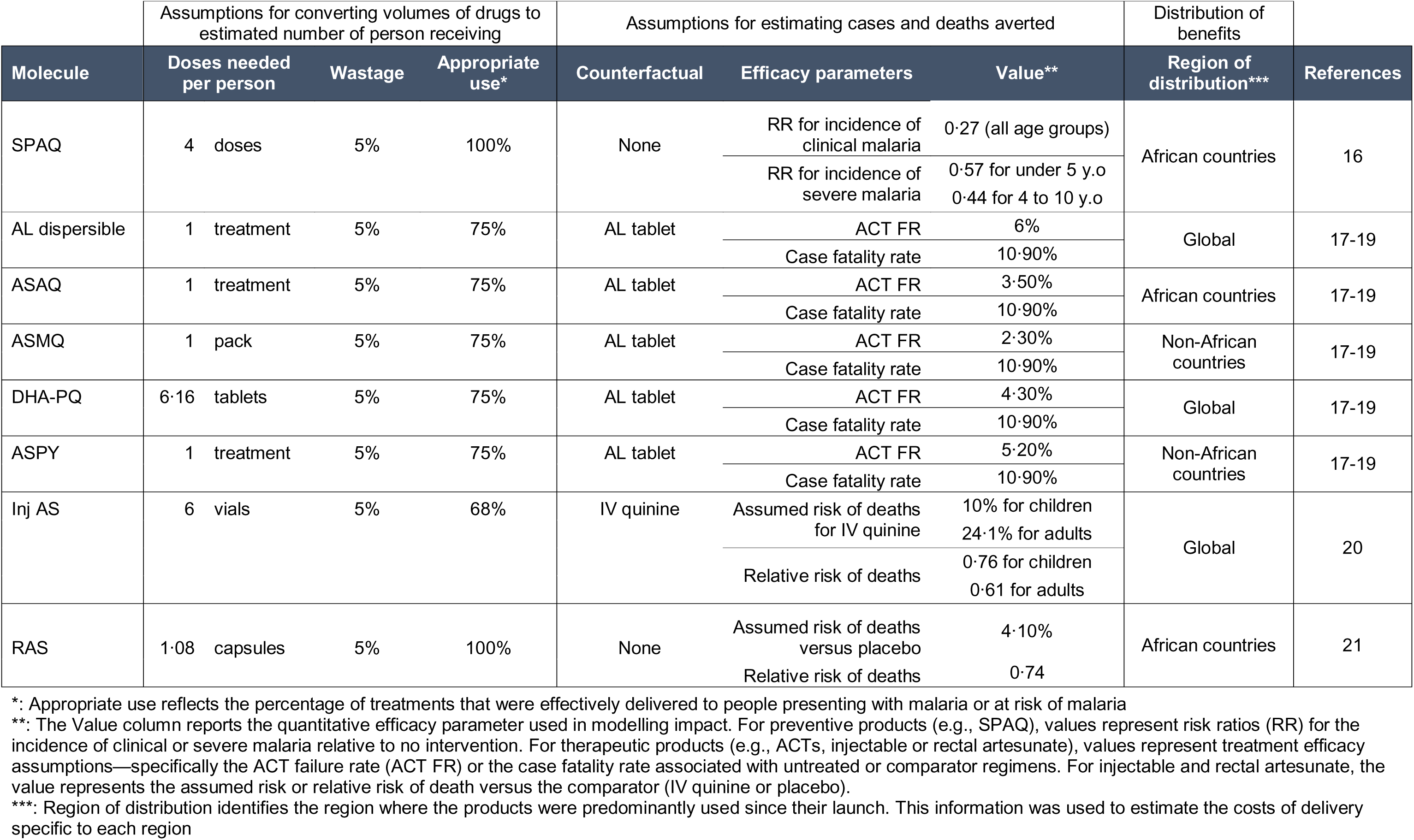
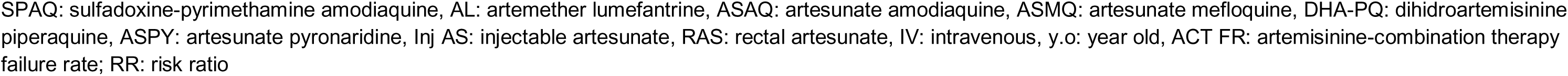
List of assumptions used to estimate the number of cases averted, severe cases averted, and deaths averted by product developed.

### Step 3: estimating and monetizing DALYs

The model calculates the number of DALYs averted based on the estimated number of lives saved, and uncomplicated and severe cases averted. Neurological sequelae were the only disabilities included, as they represent the most clinically relevant long-term outcome of severe malaria. However, their contribution to total DALYs is small relative to the burden arising from premature mortality (YLL).^21,22^ Assumptions for average life-expectancy among children aged 1-4 in high-burden populations was based on Pfeil et al.^23^ The adjustments for neurological sequelae were based on Lubell et al and Okebe et al.^21,22^

Value of Statistical Life (VSL) was used to translate the DALYs into economic terms. Recognizing that VSL can vary significantly with income levels, we adjusted the base U.S. VSL to reflect the economic context of each country in our analysis (supplementary material B). This adjustment uses a formula that incorporates the country’s income level and the income elasticity of VSL. This approach is consistent with the methodology employed by others and is aligned with Reference Case Guidelines for Benefit-Cost Analysis in Global Health and Development (supplementary material C).^2,5^ This allowed to estimate VSL for each country and subsequently determine the overall economic value of the health gains achieved.

### Step 4: Estimating the costs of delivery

To estimate the cost of delivery, we identified the 15 countries with the highest malaria burden in the regions in which MMV’s antimalarial medicines are marketed for every year of the time period since the launch of the first product in 2009. As this subset of countries changes from year to year, a total of 36 individual countries were included in the analysis, representing between 77% and 82% of global malaria cases annually.^24^ This dynamic selection of the 15 highest-burden countries in any given year ensures the analysis remains relevant and reflects the shifting geographical distribution of malaria.

We use the WHO Choice Costing tool and Global Fund pricing data to estimate country specific health care delivery costs and costs of health commodities respectively.^25,26^ We then multiplied country specific delivery costs, rapid diagnostic test costs, and treatment costs with the number of doses available to a country in a given year to arrive at an estimate of the total health system costs, sensitive to country-specific factors. ^25,26^

### Sensitivity analysis

The baseline analysis assumed a 3% discount rate, full attribution of investment returns to the PDP, an income elasticity of 1·2 for VSL, and a midpoint estimate of DALYs averted based on product-specific mortality reduction assumptions. These assumptions form the reference case against which all sensitivity analyses were compared.

We conducted a probabilistic sensitivity analysis to assess how uncertainty in treatment costs and healthcare delivery costs impacts our results. These parameters were modelled using a beta-PERT distribution for probabilistic sampling, chosen to reflect realistic ranges and potential skewness. Bounds for treatment costs were derived from empirical data, while those for healthcare delivery costs were based on WHO-CHOICE estimates, with upper and lower limits informed by their standard deviation.^26^ We generated 1,000 independent parameter sets

via Monte Carlo simulation and calculated 95% uncertainty intervals from the 2·5th and 97·5th percentiles of the resulting distribution. We calculated t-tests and confidence intervals to quantify the uncertainty around the mean IRR and BCR estimates. Additionally, we conducted a one-way sensitivity analysis to examine the influence of individual assumptions on the base case estimates. Specifically, we adjusted the discount rate to 2% and 5%, reflecting commonly used lower and upper bounds in global health economic evaluations. The income elasticity of the VSL was varied to 1·5, a more conservative estimate frequently cited in the literature for transferring VSL across different income settings.^27^ Additionally, we modified the estimated number of DALYs averted based on product-specific clinical trial efficacy data, encompassing both low and high mortality impact scenarios.

The model was built using the statistical software R. The code and data are accessible on GitHub repository (https://github.com/paulachristen/mmv_ror/).

### Role of funding source

The study received no funding.

## RESULTS

A total of US$ 2·3 billion (2023 prices) was invested into MMV during the period 2000 to 2023 (Source MMV). During that period, MMV has been involved in the clinical development of over 45 products, with 16 products being commercially available and 12 generating sales by 2023 (supplementary material A).^28^ The investment received, number of deaths averted, DALYs averted, monetized DALYs and cost of product delivery are shown in Table 3.

**Table 3:** Yearly investments, deaths averted, DALYs averted, monetized DALYs and Cost of delivery – 2000 to 2023 in US$2023.

| Year | # of MMV-supported products* | Investment in 2023 USD | Deaths averted | DALYs averted | Monetized DALYs | Cost of delivery |
| --- | --- | --- | --- | --- | --- | --- |
| 2000 |  | 29'852'923 | 0 | 0 | 0 | 0 |
| 2001 |  | 50'108'598 | 0 | 0 | 0 | 0 |
| 2002 |  | 32'524'927 | 0 | 0 | 0 | 0 |
| 2003 |  | 61'183'848 | 0 | 0 | 0 | 0 |
| 2004 |  | 76'280'272 | 0 | 0 | 0 | 0 |
| 2005 |  | 114'095'421 | 0 | 0 | 0 | 0 |
| 2006 |  | 71'055'329 | 0 | 0 | 0 | 0 |
| 2007 |  | 173'925'704 | 0 | 0 | 0 | 0 |
| 2008 | 1 | 118'242'843 | 0 | 0 | 0 | 0 |
| 2009 |  | 85'861'725 | 0 | 0 | 0 | 0 |
| 2010 | 1 | 116'044'460 | 0 | 0 | 0 | 0 |
| 2011 | 1 | 128'294'997 | 0 | 0 | 0 | 1'245'178'150 |
| 2012 | 1 | 103'748'180 | 19'525 | 1'062'171 | 3'045'809'911 | 1'123'265'199 |
| 2013 |  | 106'873'147 | 21'339 | 1'160'843 | 3'294'351'524 | 986'884'769 |
| 2014 | 1 | 126'230'106 | 85'357 | 4'643'372 | 13'039'551'031 | 1'081'783'948 |
| 2015 | 3 | 135'116'139 | 67'167 | 3'659'388 | 10'128'422'953 | 887'511'105 |
| 2016 |  | 97'182'201 | 106'688 | 5'817'429 | 16'159'551'540 | 1'023'726'289 |
| 2017 | 1 | 120'315'216 | 122'252 | 6'669'136 | 18'557'295'136 | 1'441'757'632 |
| 2018 | 2 | 89'798'807 | 139'248 | 7'600'193 | 21'316'427'266 | 1'294'064'863 |
| 2019 | 1 | 93'514'565 | 145'246 | 7'945'798 | 22'487'299'754 | 1'316'807'567 |
| 2020 |  | 108'765'449 | 193'922 | 10'594'756 | 29'842'567'014 | 2'079'103'652 |
| 2021 |  | 100'845'446 | 240'314 | 13'172'803 | 37'559'448'069 | 2'037'310'875 |
| 2022 |  | 81'301'805 | 235'464 | 12'865'771 | 36'563'924'610 | 1'922'173'152 |
| 2023 |  | 77'988'063 | 214'947 | 11'737'376 | 33'277'928'600 | 2'399'686'171 |
| **Total** | **12** | **2'299'150'173** | **1'591'470** | **86'929'036** | **10'219'690'725** | **784'968'890** |
| *: indicates the year in which MMV-Supported products were either launched or entered MMV's portfolio<br>DALYs: disability adjusted life years |  |  |  |  |  |  |

Table 3 illustrates the lag time between investment in product development and realization of the health gains. The first deaths averted by a PDP-supported product appeared 12 years after the creation of the PDP and initial investments in antimalarial drug development. Health gains from supported products increased steadily from 2011 to 2021. The reduction in deaths averted in 2022 and 2023 was due to delayed health gains following a spike of procurement of drugs for severe malaria in 2020 and 2021.

Our base case analysis finds an IRR of 52·13% and a BCR of 12·99 (Table 4). In other words, every dollar spent on MMV’s R&D delivers an additional return of 52 cents every year, for ever, and for every $1 invested in MMV for the period 2000 to 2023, $12·99 is recouped in monetary benefits. Varying the discount rate has minimal impact on IRR estimates, ranging from 52·30% to 51·27%, and BCR estimates ranging from 12·31 to 14·07 for the period 2000 to 2023. The only parameter to show any marked change to the base case results was when we tested an elasticity of 1·5. The IRR decreased to 33·86% and the BCR to 3·84 for the study period, representing a 35% and 70% decrease compared to the base case, respectively. The IRR varies from 48·06% for the low DALY scenario to 54·49% in the high DALY scenario, while BCR varied from 8·98 to 16·32 for the study period, respectively.

**Table 4:**
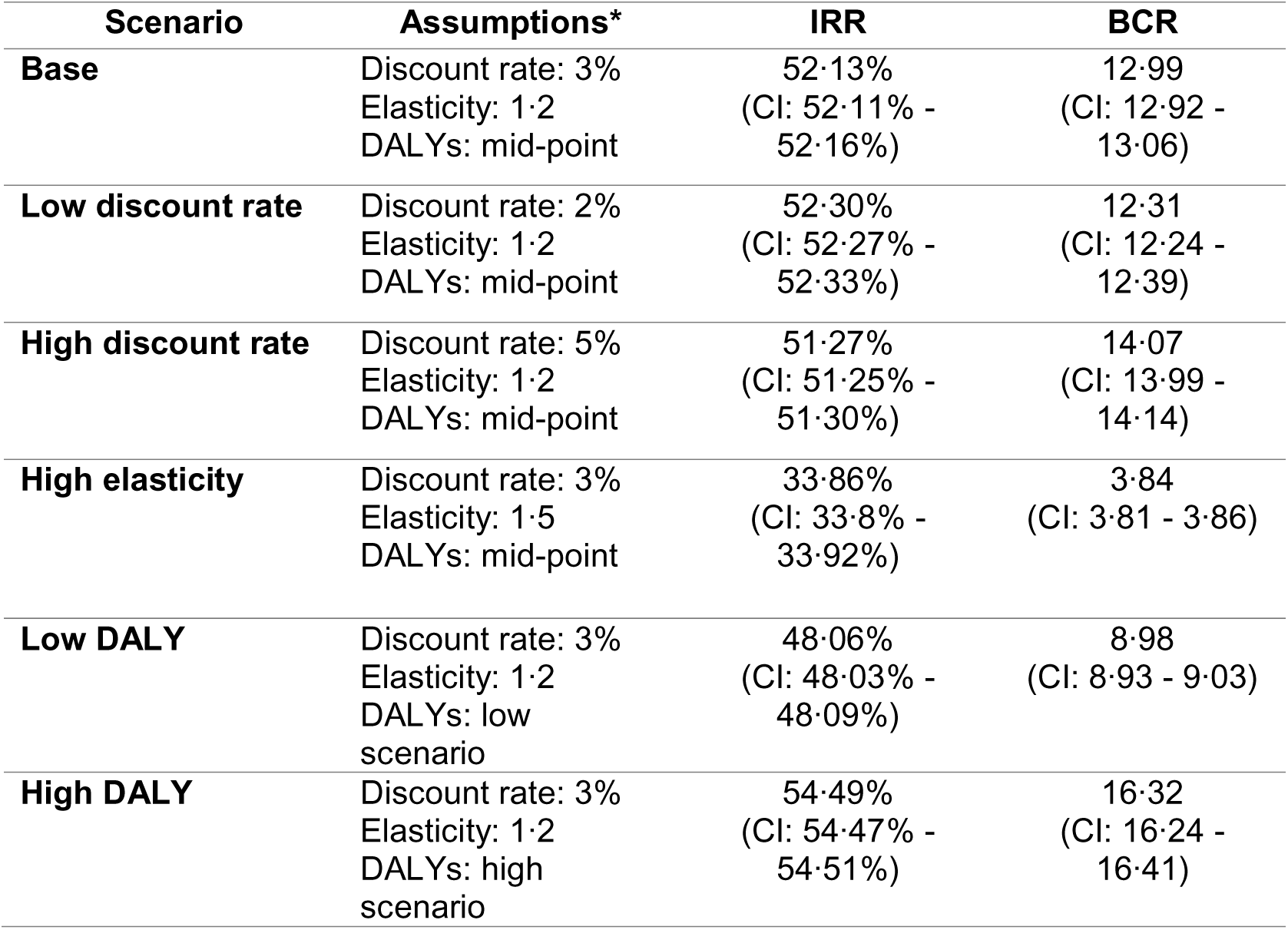
Internal Rate of Return and Benefit-Costs ratio for various scenarios.

| Scenario | Assumptions* | IRR | BCR |
| --- | --- | --- | --- |
| **Base** | Discount rate: 3%<br>Elasticity: 1.2<br>DALYs: mid-point | 52.13%<br>(CI: 52.11% - 52.16%) | 12.99<br>(CI: 12.92 - 13.06) |
| **Low discount rate** | Discount rate: 2%<br>Elasticity: 1.2<br>DALYs: mid-point | 52.30%<br>(CI: 52.27% - 52.33%) | 12.31<br>(CI: 12.24 - 12.39) |
| **High discount rate** | Discount rate: 5%<br>Elasticity: 1.2<br>DALYs: mid-point | 51.27%<br>(CI: 51.25% - 51.30%) | 14.07<br>(CI: 13.99 - 14.14) |
| **High elasticity** | Discount rate: 3%<br>Elasticity: 1.5<br>DALYs: mid-point | 33.86%<br>(CI: 33.8% - 33.92%) | 3.84<br>(CI: 3.81 - 3.86) |
| **Low DALY** | Discount rate: 3%<br>Elasticity: 1.2<br>DALYs: low scenario | 48.06%<br>(CI: 48.03% - 48.09%) | 8.98<br>(CI: 8.93 - 9.03) |
| **High DALY** | Discount rate: 3%<br>Elasticity: 1.2<br>DALYs: high scenario | 54.49%<br>(CI: 54.47% - 54.51%) | 16.32<br>(CI: 16.24 - 16.41) |

## DISCUSSION

PDPs have emerged as a pivotal strategy for donors to support research for neglected diseases of poverty.^9^ To our knowledge, this study is the first systematic bottom-up attempt to estimate the return on investment from a single PDP. Previous mixed methods approaches have focused on describing the reach of PDPs but not quantified their impact in monetary terms across an entire portfolio.^7^ Rather, examples of specific innovations supported by PDPs have been estimated.^11^ Furthermore, our analysis is rare as we are able to base our findings on retrospective data of confirmed volumes of drugs distribution that were routinely collected by MMV, rather than the more common approach of projecting hypothetical product use into the future. With an IRR of 52·13% (CI: 52·11% – 52·16%) our findings indicate that for every dollar invested in the PDP to date, a return of 52 cent is expected every year into the future. In addition, a BCR of 12·99 (CI: 12·92 – 13·06) indicates that for every $1 invested, society received $13 worth of health benefits in return for the period 2000 to 2023. Both metrics show this PDP was a highly worthwhile investment.

While both the BCR and IRR capture investment efficiency, they provide complementary perspectives. The IRR reflects the annualised rate of return, incorporating the timing of costs and benefits, and is therefore sensitive to the pace at which health gains are realised. The BCR, in contrast, expresses the total social value generated per dollar invested, irrespective of time. In contexts where benefits accrue gradually, as with most product development pipelines, the BCR can appear high even if returns are realised over decades. By contrast, the IRR is more informative for comparing investments with differing time horizons or cash flow patterns. Both metrics depend on the magnitude of health gains, but the IRR is particularly sensitive to when those gains occur. If malaria burden were to decline substantially in the future due to wider control measures, the realised BCR would remain unchanged for the period analysed (2000–2023), but any prospective IRR incorporating future projections would decline as fewer DALYs could be averted by the same interventions. Conversely, if disease burden increased or PDP-supported products expanded coverage, the IRR for future periods would rise. Hence, while our retrospective analysis provides a conservative estimate of realised returns, future ROI estimates will necessarily evolve with changes in disease epidemiology and product uptake.

It is useful to interpret these figures and situate them in the wider literature, however comparing return on investment calculations is challenging due to different assumptions, across different timelines, with a range of approaches to discounting financial and health inputs and outputs.^1^ Such variable methodologies often make direct comparations misleading. Even with all the caveats and cautions, there is value in drawing upon the wider literature. Our findings are in line with previous studies which suggest that investing in scientific research yields returns that can largely exceed the initial costs. One highly influential UK report suggested that every £1 of public investment medical research generated an additional 25 UK pence annually in health gains and GDP benefits. ^29^ While the authors acknowledged the challenges in directly comparing this return to other areas of public spending, they noted that it significantly exceeds the typical 6-8% annual return expected by the UK government on public investments.^29^ In 2024, a comprehensive study compared the estimated long-term health impact of new products for neglected diseases (launched since 2000 or projected to be launched before 2040) against the total global investment required for their development from 1994 to 2040·^5^ This analysis included global funding across the full range of neglected diseases, including new vaccines for bacterial pneumonia and meningitis, cholera, rotavirus, malaria, typhoid, and tuberculosis (TB), improved diagnostics for HIV and TB, improved drugs for HIV, TB and malaria, and malaria vector control. They estimated that every $1 invested in neglected disease R&D generated a ROI of $405, mainly due to the societal value of lives saved, timelines, and scope. In 2025 the same group estimated the impact of UK’s £3·05 billion funding for neglected disease R&D would generate a global social returns worth £1·39 trillion for the period 2000 to 2040.^30^

The methodology we adopted varies in two important ways from previous analyses. First, we did not forecast the health gains of existing and future products past 2023. In other words, we only calculated the IRR and BCR for the commercially available products that have been used by 2023, and we do not account for the future health gains of products already launched or currently under development. Many return on investment analyses are often framed from an advocacy perspective where the hypothetical benefits from a new intervention or strategy are modelled by forecasting potential future gains. For example, the projected return on investment of RTS,S/AS01 malaria vaccine varied from 0·42 to 2·30 for the period 2021 to 2030, depending on the assumptions used for the calculations.^4^ However, the actual return on investment for malaria vaccines, based on real life data, has not yet been evaluated. Second, our case study differs from previous analyses by estimating the return on investment for the entire portfolio of products supported by a single PDP. In contrast, earlier studies have focused on individual products or interventions,^4,22,23^ specific diseases,^6,12^ or aggregated all contributors involved in the new product development.^5^ Our case study demonstrates how standard metrics can be effectively applied to assess the returns generated by the PDP model.

Despite recognized limitations in disability weights, age weighting, and cultural context, we used DALYs, the most widely accepted and policy-relevant metric for global health evaluations to ensure comparability with previous global health investment analyses. ^1,2, 5, 6^ Sensitivity analyses further highlighted that assumptions about elasticity had the greatest influence on the estimated returns. Elasticity reflects how strongly health benefits respond to changes in product coverage and uptake, in other words how much impact an increase in access or use has on overall health outcomes. Because of this relationship, even small adjustments in elasticity values lead to disproportionately large shifts in the estimated ROI.

It is also important to note that most PDP funding remains concentrated on research and development activities, while efforts related to product access and delivery continue to receive limited support. This imbalance constrains the realisation of full health and economic returns, as delays in ensuring widespread access slow the accrual of benefits. If access-related components - such as registration, supply chain strengthening, and health system integration - were adequately funded, a larger proportion of the target population could benefit earlier. In such scenarios, returns on investment would likely begin sooner than the 12-year average lag observed in our analysis and yield higher overall BCR and IRR values.

A key feature of the PDP model is the ability to pool funds across different donors and de-risk the development of unsuccessful projects.^11^ This approach avoids the need for funders to ‘pick winners’ at the individual product line.^9^ PDPs receive investment either as restricted grants – designated for specific programmes or objectives as defined by the donor – or as unrestricted funding, which allows them to allocate resources flexibly across operational priorities. PDPs engage in all stages of the product development cycle, from early-stage research through to regulatory approval, often advancing products more rapidly and cost-effectively than either academic institutions or private sector entities. Unrestricted funding provides PDPs the flexibility to reallocate resources efficiently across projects based on progress toward defined milestones and target product profiles.^7^ PDPs adopt a portfolio approach across an entire product class (such as antimalarials) and can make data-driven decisions to advance or discontinue candidates according to scientific outcomes. Furthermore, increased coordination and pooling of donor resources serve to reduce the risks associated with developing products that may not be commercially lucrative. This approach incentivises pharmaceutical industry engagement and helps to address market failures. By convening a diverse set of partners and technical expertise, PDPs can accelerate the timeline from discovery to clinical evaluation, licensure and the delivery of affordable, safe, effective and high-quality health products.

### Limitations

Our approach has its limitations. First, a VSL approach was used to translate the DALYs into economic benefits. We appreciate that this represents population-level willingness to pay to reduce the risk of death, not the value of an individual life. It is common and appropriate to adjust the US reference VSL to reflect income levels of other countries, especially in global health economic analysis, as recommended by the OECD, World Bank etc. It helps ensure that benefit estimates are contextually realistic and not overstated to allow for fairer comparisons across countries with different income levels. However, we recognise that the debate continues on the ethical and equity implications of this approach.

An additional limitation concerns the estimation of delivery costs. Comprehensive data on the cost of healthcare delivery of antimalarial drugs have not been consistently available across all countries since 2000. Consequently, our analysis focused on the 15 countries with the highest annual malaria burden and where MMV-supported products were deployed. While this approach likely underestimates the total cost of delivery, these countries accounted for at least 77% of global malaria cases in any given year, thus capturing the majority of the disease burden. Moreover, the calculated IRR and BCR pertain specifically to the portfolio of products developed with PDP support.

Finally, the assessment of the potential health impact resulting from the delivery and distribution of the PDP-supported products relies on routinely available data to estimate the proportion of malaria treatments administered to patients with malaria. As with all modelling efforts, a key area of uncertainty lies in the extent to which health system inefficiencies may hinder the effective translation of distributed treatments into actual coverage and clinical impact.

## Conclusion

Our aim was to adapt existing methodologies for estimating the return on investment from global health research using malaria medicines as a case study. This study has also enabled us to highlight some of the methodological and data challenges which could be explored in future research, allowing for us to investigate the extent our findings are generalisable. In a world where there are likely to be far fewer donor resources for development, it is important that the full impacts of different investments are quantified. It is hoped that this study will help inform approaches on the evaluation of R&D investments and will add to the academic literature and evidence base on the potential cost-benefit of development interventions and programmes, including research. Also, from an empirical perspective - even in the context of some very conservative assumptions - our analysis confirms the value of investment in this area.

## Author’s contributions

CvD and OBH developed the model estimating the lives saved and DALYs, with guidance and inputs from CA. PC calculated the IRR and BCR with guidance and inputs from LC and JM. PCP, AC, JM and LC conceptualized the paper and assembled the first draft. All authors reviewed the results and contributed to the manuscript.

## Declaration of interests

PC is an employee of Contemplativus in Actione Ltd. CA is an employee of MMV. JAM is an employee of FCDO. OBH is an employee of CEPA and acted as a consultant for this work. CvD is an employee of CEPA and acted as a consultant for this work. LC is an employee of London School of Economics and a Wellcome Trust Career Development Awardee [Grant No. 317468/Z/24/Z]. Authors declare no conflict of interest. The views reflected in this paper are entirely those of the named authors and do not reflect UK or any government policy.

## Data sharing statement

This study did not involve the collection of individual participant data. A data dictionary defining all fields in the study dataset will be made publicly available via GitHub to support reproducibility. No deidentified or identifiable participant data are included.

Additional study materials, including the statistical analysis code and a README describing the structure and use of the dataset, will also be available.

These materials are already accessible and will remain available indefinitely from the following URL: https://github.com/paulachristen/mmv_ror/

All shared resources will be openly accessible and may be used by any researcher without restriction, in accordance with principles of reproducible research. No prior approval or data access agreement will be required.

## Data Availability

All data produced are available online at

https://github.com/paulachristen/mmv_ror/

## Acknowledgements

Authors wish to thank George Jagoe for his guidance on this work throughout the whole project.

## Declaration of generative AI and AI-assisted technologies in the writing process

During the preparation of this work the authors used ChatGPT-4o in order to improve text clarity. After using this tool, the authors reviewed and edited the content as needed and take full responsibility for the content of the publication.

## Notes

### Competing Interest Statement

The authors have declared no competing interest.

### Funding Statement

This study did not receive any funding

### Summary of Updates

Amendments made based on reviewer feedback

