## Supplement for "Measuring the Impact of Global R&D Investment in Product Development Partnerships (PDPs): A Case Study on Return on Investment in Antimalarial Drug Development"

**Supplementary material**

A. **MMV-supported product introduction timeline**

**Figure 1. MMV-supported product introduction timeline.**

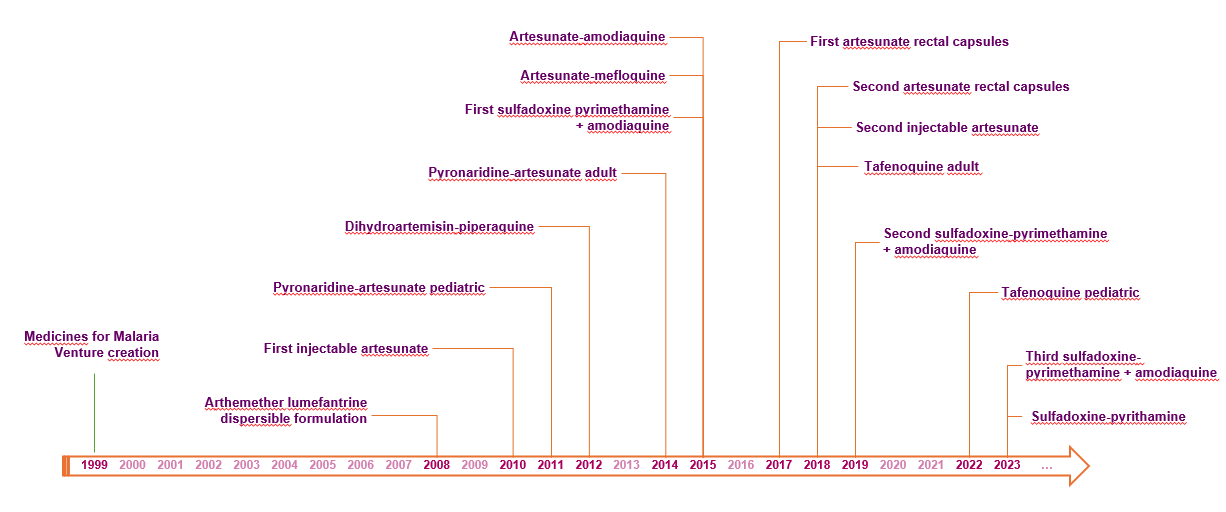

B. **Estimated cost of treatment delivery per country**

**Table 1. Estimated cost of treatment delivery per country in 2023 USD.**

| **Country** | **Health Systems Costs (in 2023 USD)** |
| --- | --- |
| Nigeria | 7,656,603,214 |
| Angola | 1,641,952,384 |
| Congo, Dem. Rep. | 1,201,083,978 |
| Uganda | 1,075,036,423 |
| Cote d'Ivoire | 863,706,812 |
| Cameroon | 845,710,744 |
| Ghana | 759,311,127 |
| Tanzania | 683,894,118 |
| Burkina Faso | 663,118,712 |
| Mali | 637,164,237 |
| Mozambique | 570,645,850 |
| India | 482,201,483 |
| Niger | 418,885,607 |
| Ethiopia | 409,746,731 |
| Benin | 344,264,354 |
| Guinea | 188,470,770 |
| Rwanda | 142,332,525 |
| Malawi | 138,275,622 |
| Madagascar | 74,618,016 |
| Pakistan | 13,338,193 |
| Indonesia | 10,335,415 |
| Papua New Guinea | 9,377,095 |
| Yemen | 6,653,579 |
| Venezuela (Bolivarian Republic of) | 6,509,114 |
| Colombia | 4,247,033 |
| Myanmar | 1,523,374 |
| Solomon Islands | 1,038,671 |
| Afghanistan | 1,000,978 |
| Brazil | 933,137 |
| Peru | 766,276 |
| Cambodia | 380,348 |
| Guyana | 167,431 |
| Nicaragua | 122,906 |
| Haiti | 89,277 |
| Bangladesh | 67,838 |

C. **Estimation of the Value of a Statistical Life Year (VSLY)**

To estimate a country-level Value of a Statistical Life Year (VSLY), we first calculated a DALY-weighted global average income figure. This process involved the following steps:

1. **Weighting DALYs by national income:**
   We summed total discounted DALYs by country (discounted at 3%, with a base year of 2023) and calculated each country’s proportional share of the total DALYs (denoted as DALYs_share).
2. **Income adjustment using GNI per capita:**
   We then multiplied each country’s DALYs_share by its gross national income (GNI) per capita, adjusted for purchasing power parity (Atlas method), using 2023 data from the World Bank. If a country’s GNI per capita was not available for 2023, we used the most recent available figure.
3. **Calculating the DALY-weighted average income:**
   This yielded a DALY-weighted income measure for each country (GNI_DALYs_weighted).
4. **Estimating the Value of a Statistical Life (VSL):**
   Using the Gates Reference Case and CEPI guidance, we applied an income elasticity of 1.2 to estimate a DALY-weighted global VSL. We used a U.S. reference VSL of $12.3 million and a U.S. GNI per capita of $80,450 (World Bank, 2023):

VSL = 12,300,000 × (total_GNI_DALYs_weighted / 80,450)^1.2

5. Finally, we converted the VSL to a Value of a Statistical Life Year (VSLY), assuming that a VSL corresponds to 40 life years and applying a discount rate of 3%:

years = 40

discount_rate = 0.03

inverse_discount_rate = 1 / (1 + discount_rate)

VSLY = [VSL × (1 - inverse_discount_rate)] / [1 - (inverse_discount_rate^years)] × (1 + discount_rate)

This process allowed us to derive a globally relevant VSLY that reflects both differences in income across countries and the distribution of the health burden addressed by the intervention.

**WORKED EXAMPLE: VSLY ESTIMATION STEP-BY-STEP**

To illustrate the estimation of VSLY, we present a worked example using three high-burden malaria countries with 2023 data. This example demonstrates how absolute disease burden flows through each step of the calculation.

**SAMPLE DATA (2023 Malaria Burden)**

| Country | Year | Malaria Cases | GNI per capita (PPP) |
| --- | --- | --- | --- |
| Nigeria | 2023 | 68,136,453 | $1,880 |
| Democratic Republic of the Congo | 2023 | 33,140,568 | $630 |
| Uganda | 2023 | 12,572,518 | $970 |

**Total malaria cases: 113,849,539**

**STEP 1: Calculate each country's share of malaria burden**

We calculate the proportion of global malaria burden for each country:

burden_share = country malaria cases / total global malaria cases

Results:

- Nigeria: 68,136,453 / 113,849,539 = 0.5985 (59.8%)
- DRC: 33,140,568 / 113,849,539 = 0.2911 (29.1%)
- Uganda: 12,572,518 / 113,849,539 = 0.1104 (11.0%)

Note: The burden_share is a PROPORTION of global malaria cases. The sum of all proportions equals 1.00 (100%).

**STEP 2: Weight GNI per capita by malaria burden distribution**

We multiply each country's GNI per capita by its burden_share:

GNI_burden_weighted = burden_share × GNI per capita

Results:

| Country | GNI per capita | burden_share | GNI_burden_weighted |
| --- | --- | --- | --- |
| Nigeria | $1,880 | 0.5985 | $1,125.14 |
| DRC | $630 | 0.2911 | $183.39 |
| Uganda | $970 | 0.1104 | $107.12 |

This step shows how countries with higher malaria burden contribute more to the weighted average. Nigeria, with 59.8% of cases, contributes $1,125.14 to the weighted average despite not having the highest GNI per capita.

**STEP 3: Sum GNI_burden_weighted across all countries**

We sum the burden-weighted GNI values to obtain a single average:

total_GNI_burden_weighted = Σ(GNI_burden_weighted) total_GNI_burden_weighted = $1,125.14 + $183.39 + $107.12 = $1,415.64

This represents the average GNI per capita, weighted by malaria burden. This weighted average ($1,415.64) is much closer to Nigeria's GNI per capita ($1,880) than to a simple arithmetic mean of the three countries ($1,160), because Nigeria bears the largest share of the malaria burden.

**STEP 4: Calculate Value of Statistical Life (VSL)**

We use the income elasticity approach with the following formula:

VSL = VSL_US × (total_GNI_burden_weighted / GNI_US)^elasticity

Inputs:

- U.S. VSL: $12,300,000
- U.S. GNI per capita: $80,450
- Weighted GNI per capita: $1,415.64
- Income elasticity: 1.2

Calculation: VSL = $12,300,000 × ($1,415.64 / $80,450)^1.2 VSL = $12,300,000 × (0.017597)^1.2 VSL = $12,300,000 × 0.007844 VSL = $96,476

The income elasticity of 1.2 means that VSL increases by 1.2% for every 1% increase in income. This super-elastic relationship reflects empirical findings that willingness to pay for mortality risk reductions increases more than proportionally with income.

**STEP 5: Convert VSL to VSLY (Value of Statistical Life Year)**

We use the annuity formula to convert the lump sum VSL into an equivalent annual value:

VSLY = [VSL × (1 - 1/(1+r))] / [1 - (1/(1+r))^n] × (1+r)

where r = discount rate and n = life years

Inputs:

- Years: 40
- Discount rate: 0.03 (3%)
- Inverse discount rate (1/1.03): 0.970874

Calculation steps:

Step 5a: Calculate numerator Numerator = VSL × (1 - 1/1.03) Numerator = $96,476 × 0.029126 Numerator = $2,810

Step 5b: Calculate denominator Denominator = 1 - (1/1.03)^40 Denominator = 1 - 0.306557 Denominator = 0.693443

Step 5c: Divide and adjust VSLY = ($2,810 / 0.693443) × 1.03 VSLY = $4,052 × 1.03 VSLY = $4,174

**SUMMARY OF RESULTS**

| Metric | Value |
| --- | --- |
| Total malaria cases | 113,849,539 |
| Burden-weighted GNI per capita | $1,415.64 |
| Value of Statistical Life (VSL) | $96,476 |
| Value of Statistical Life Year (VSLY) | $4,174 |

This VSLY value would be used to monetize DALYs averted in our analysis. For example, if an intervention averts 1,000 DALYs:

Monetized benefit = 1,000 × $4,174 = $4,174,000
